# Risk Assessment of COVID-19 Airborne Infection During Hybrid Learning

**DOI:** 10.1101/2020.09.24.20200782

**Authors:** Luis A. Anchordoqui, Eugene M. Chudnovsky, Thomas C. Paul

## Abstract

Converging lines of evidence seem to indicate that SARS-CoV-2, the novel coronavirus responsible for the COVID-19 pandemic, can be transmitted from person-to-person via aerosols that waft through the air and accumulate over time. The airborne nature of the virus could be a threat in indoor spaces in general and in particular for in-class education. We provide an assessment of the risk of SARS-CoV-2 infection during a 7-hour school day in elementary schools. We show that existing data are insufficient to establish a low (below 1%) probability of infection with high accuracy. The use of facemasks and social distancing could significantly decrease this risk.

---

The coronavirus disease 2019 (COVID-19) caused by the severe acute respiratory syndrome coronavirus 2 (SARS-CoV-2) is an ongoing global health emergency [1–3]. SARS-CoV-2 is only the third strain of coronavirus known to frequently cause severe symptoms in humans. The other two strains cause the Middle East respiratory syndrome (MERS-CoV) and the severe acute respiratory syndrome (SARS-CoV). The epicenter of the COVID-19 outbreak is in the city of Wuhan in Hubei Province of central China [4]. The respiratory disease has spread rapidly across six continents and therefore the outbreak has been declared a global pandemic by the World Health Organization.

A pioneering study conducted in the city of Shenzhen near Hong Kong has provided the first concrete evidence for human-to-human transmission of SARS-CoV-2 [5]. Indeed, SARS-CoV-2 can spread from person-to-person in an efficient and sustained way by coughing and sneezing. Moreover, this coronavirus can spread from seemingly healthy carriers or people who had not yet developed symptoms [6].

Experiment suggests that SARS-CoV-2 may have the potential to be transmitted through aerosols [7–11]. Actually, laboratory-generated aerosols with SARS-CoV-2 were found to keep a replicable virus in cell culture throughout the 3 hours of aerosol testing [12]. The estimated lifetime of the aerosolized virus is about 2 hr. Stronger and stronger evidence suggest that the respiratory particles emitted during a sneeze or cough are initially transported as a turbulent cloud that consists of hot and moist exhaled air and mucosalivary filaments [13, 14]. Numerical simulations and analytic studies show that aerosols and small droplets trapped inside the turbulent puff cloud could propagate up to 7 or 8 m [15–18]. Besides, once the cloud slows down sufficiently, and its coherence is lost, the eventual spreading of the infected aerosols becomes dependent on the ambient air currents and turbulence. In particular, convective airflow influence the distribution of viral particles in indoor spaces, cultivating a health threat from COVID-19 airborne infection [19].

The airborne nature of SARS-CoV-2 imply that hybrid learning models being considered by school districts for reopening in the Fall could pose a community transmission risk. In this Letter we investigate the risk of SARS-CoV-2 infection during a 7-hour school day in elementary schools.

A contagious student will shed virions into the classroom by breathing, talking, coughing, and sneezing. Infectious aerosols are separated by large volumes of uninfected air. The aerosolized number density (a.k.a. concentration) of virions *n* in the classroom is governed by a balance equation

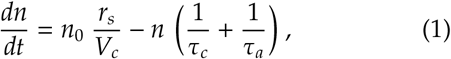

where *n*_0_ is the concentration of virions in saliva (or viral load) that is emitted by the source in aerosol form at a rate *r*_*s*_, *V*_*c*_ ∼ 250 m^3^ is the typical volume of a classroom in the U.S. [20], *τ*_*a*_ ∼2 hr is the viral decay time in aerosol form [12, 21], *τ*_*c*_ *= V*_*c*_/*r*_*c*_ is the air cycle-time in the classroom, and *r*_*c*_ is the rate at which air is removed from the classroom (or filtered locally) [22]. The viral load in saliva has a large uncertainty varying in the range 10^3^ < *n*_0_/mL < 10^11^ [22–26]. A recent comprehensive study has shown that (asymptomatic) children can carry viral loads exceeding those of very sick adult patients cared for in intensive care units [28]. This could make them ideal silent spreaders of COVID-19. Therefore, in our calculations we conservatively adopt the 95th percentile of the log normal distribution fitted to the data in [26], which is an order of magnitude below the estimate of [22].

The emission aerosol rate also varies over a long range and actually depends on the respiratory process. At a typical rate, a student will breathe out a volume of aerosols per unit time of *r*_*s*_ ∼ 120 pL/min [26]. This average value for *r*_*s*_ ∼ is also an order of magnitude smaller than the estimate of [22]. The range for the emission rate of aerosols while breathing and talking is shown in Fig. 1 together with the cough and sneeze aerosol volumes. In our study we consider a classroom ventilation system varying in the range 1 < *r*_*c*_/(m^3^ min^−1^) < 10 ~ 350 cfm. The concentration is found to be

**FIG. 1:**
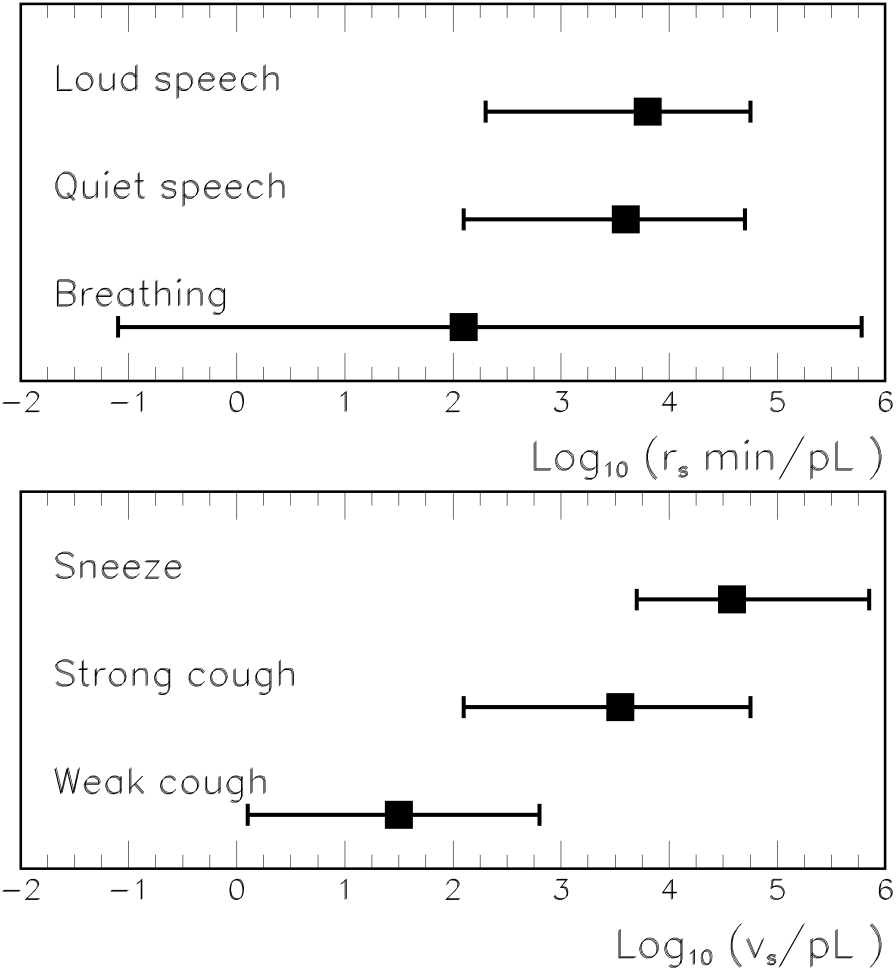
Box-whisker chart of the emission aerosol rate *r*_*s*_ and the total aerosol volume v_*s*_ that are expelled by a source *s*.

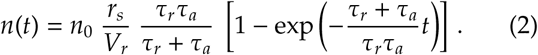

For school day, the exposure time is *t =* 7 hr, yielding 0.99 < *n*/m^3^ < 3.87.

To determine the infection probability we adopt the traditional Wells-Riley risk model [31, 32]. The total expected number of virions in given dose is estimated to be *d = nr*_*b*_*t*, where *r*_*b*_∼ 10 L/min is the breathing rate. The actual integer number of virions deposited in the dose would follow a Poisson probability distribution. If the deposition of a single virion can initiate infection, the risk of infection is equal to 1 − *e*^−*d*^. The term *e*^−*d*^ is the Poisson probability that no virions are contained in the dose, whereas the complement is the probability that one or more virions are deposited in the dose, in which case infection is assumed.

If instead *k* organisms are required to infect, the probability of developing an infection can be generalized to

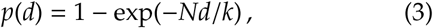

where *N* is the number of students in the classroom [26,33]. The infection constant, *k =* 400, has been derived from a likelihood fit using data from transgenic mice infected with SARS-CoV [29]. Given the similarities between the infectious properties of SARS-CoV and SARS-CoV-2 in animal models [30], we assume here that the value of *k* fitted in [29] applies to humans as well for SARS-CoV-2. The average size of a class in the U.S. is about 20 students that will be divided into two groups for hybrid learning. Hence, we set *N =* 10 and the probability of infection for our fiducial values is in the range = 0.10 < *p* < 0.33. Given this estimate together with the very large uncertainties in the salient data (see e.g. Fig. 1 and the range of *n*_0_), we find it difficult to support any claim that the risk of exposure to SARS-CoV-2 during 7 hours in a classroom is inconsequential.

We end with some caveats and limitations of our study. The main uncertainty of this type of analysis is encoded in the exponential dependence of the infection probability on both the infection constant *k* and the number of virons *d* given by Eq. (3). We stress that even if *k* for humans was in the same ballpark as it is for mice, a very little change in *k* would result in a dramatic change for *p*. The same (of course) is true about the number of virions in the dose *d*. The exponential dependences make our previous estimate inaccurate. Even though with our educated guess of main parameters one arrives at 0.10 < *p* < 0.33, this result is largely dominated by its uncertainty and indeed the range for the probability of infection is given by 0.000000001 < *p* < 1. The only reasonable conclusion to make is that *based upon the existing data it is impossible to say how big the infection probability in a classroom would be*. It is important to stress, however, that implementing social distancing with the use of facemasks that help filtering out bio-aerosol particles would significantly reduce the risk of infection.

The theoretical and computational techniques and resources used in this research were supported by the U.S. National Science Foundation, NSF Grant PHY-1620661 (L.A.A.), and the U.S. Department of Energy, Office of Science, DOE Grant DE-FG02-93ER45487 (E.M.C.). Any opinions, findings, and conclusions or recommendations expressed in this material are those of the authors and do not necessarily reflect the views of the NSF or DOE.

## Data Availability

Not applicable

